# Midfrontal Oscillatory Alterations During Gait Imagination and Observation in Parkinson’s Disease with Freezing of Gait

**DOI:** 10.1101/2025.09.17.25335996

**Authors:** Matthew Leedom, Arturo I. Espinoza, Martina Mancini, Daniel H. Lench, Arun Singh

**Author notes:** Correspondence to: Dr. Arun Singh, Biomedical and Translational Sciences, Sanford School of Medicine, University of South Dakota, 414 E. Clark St. Vermillion, SD, 57069, USA. Declaration of Competing interests: The authors declare no competing interests.

## Abstract

**Background:** Freezing of gait (FOG) is a debilitating motor symptom of Parkinson’s disease (PD) that reflects a breakdown in both motor automaticity and cognitive control. Cognitive strategies, such as motor imagery (gait imagination, GI) and action observation (gait observation, GO) are increasingly used in rehabilitation, yet their neural underpinnings in PD remain unclear.

**Objectives:** To examine midfrontal oscillations during GI and GO in healthy controls (HC), people with PD without FOG (PDFOG–), and with FOG (PDFOG+).

**Methods:** In this study, we used electroencephalography (EEG) to examine midfrontal oscillations during GI and GO in HC (n=21), PDFOG− (n=16), and PDFOG+ (n=21). Resting-state EEG (eyes-closed and eyes-open) served as baselines.

**Results:** Across tasks, both PD groups showed elevated midfrontal theta power relative to HC, while beta oscillations differentiated PDFOG– from PDFOG+. During GO, PDFOG– displayed higher power in both lower and higher frequency bands, whereas PDFOG+ showed reduced alpha and beta power compared to PDFOG–. In GI, PDFOG+ exhibited reduced beta power compared to PDFOG–. Importantly, slowing the pace of GI or GO in PDFOG+ led to reductions in delta, theta, and beta activity, approximating patterns observed in HC.

**Conclusions:** These findings suggest that GI and GO recruit overlapping but partially distinct control mechanisms, with impaired oscillatory engagement in PDFOG+. Slowing task pace appears to normalize midfrontal rhythms, potentially by reducing cognitive-motor demands. These oscillatory signatures provide mechanistic insight into gait-related dysfunction and supports the refinement of motor imagery and action observation as targeted rehabilitation strategies for PD.

## Introduction

Freezing of gait (FOG) is among the most disabling axial symptoms of Parkinson’s disease (PD), contributing to falls, loss of independence, and diminished quality of life. Individuals with PD experience approximately 62% more falls than the general population, with FOG, postural instability, and cognitive dysfunction emerging as key contributors.^1, 2^ While pharmacological treatments and neuromodulation have offered solutions to alleviate some of the cardinal features of PD, including bradykinesia, tremor, and rigidity, their effect on axial and cognitive symptoms remains limited.^3, 4^ As a result, FOG remains a major therapeutic challenge.

The neurophysiological mechanisms underlying gait impairments in PD remain poorly understood, in part due to the complexity of the neural networks involved in gait and the unpredictable nature of FOG.^5, 6^ Gait depends not only on automatic motor programs, but also on cognitive processes such as executive control, attention, and motor planning. Disruption of these systems is strongly linked to gait dysfunction and FOG in PD.^7, 8^ Rehabilitation approaches that target these cognitive functions, including motor imagery (gait imagination, GI) and action observation (gait observation, GO), have shown promise in enhancing motor performance and reducing freezing episodes.^9–11^ GI engages internal motor planning networks, whereas GO relies on externally triggered perceptual-motor coupling, offering complementary avenues to support locomotor control.^12, 13^

Neuroimaging studies demonstrate altered cortical control of gait in people with FOG (PDFOG+), including changes in the mesencephalic locomotor region and frontal areas, compared to people with PD who do not have FOG (PDFOG–) or healthy controls.^14, 15^ GO tasks, in particular, have been shown to facilitate motor learning and improve walking in people with PD who have gait dysfunction, likely through neuroplastic changes in cortical regions.^9, 12^ Abnormal cortical oscillations also appear central to gait disruption in PD. Reduced frontal cortical theta rhythms (4-8 Hz) have been implicated in impaired cognitive control and executive functions in people with PD,^16, 17^ while elevated beta rhythms (13-30 Hz) in motor cortical-basal ganglia networks have been associated with impaired motor task execution.^18, 19^ A relationship between increased beta band oscillations in the cortico-basal ganglia network and gait-related symptoms in PD has also been established.^18–20^ Our previous research demonstrated abnormal midfrontal theta and beta oscillations in PDFOG+ compared to PDFOG– and healthy controls during cue-based lower-limb movements.^21^ However, electrophysiological mechanism underlying GI and GO tasks in the midfrontal cortical regions of people with PD and FOG (PDFOG+) and without freezing of gait (PDFOG–) remains unclear.

This study aimed to examine midfrontal oscillatory activity during GI and GO in PDFOG+, PDFOG–, and healthy controls. By focusing on theta and beta oscillations at the midfrontal cortex, we sought to characterize group-specific oscillatory patterns and determine whether manipulating task (normal vs. slow) modulates activity in any way. Clarifying these mechanisms could refine the use of GI and GO as targeted neurorehabilitation strategies to improve gait and reduce FOG, while advancing mechanistic understanding of gait dysfunction in PD.

## Material and Methods

### Participants and clinical assessments

The study included three groups of participants: 34 PDFOG+, 16 PDFOG–, and 21 healthy controls. PDFOG+ status was determined using a score greater than 1 on item 3 of the Freezing of Gait Questionnaire (FOGQ). The PDFOG+ group was further divided into two subgroups. The first subgroup (21 PDFOG+) performed the GI and GO tasks at a normal walking speed, following instructions identical to those given to the PDFOG– and healthy control groups. The second subgroup (13 PDFOG+) completed the tasks at a slower walking speed. Participants with PD were recruited from local neurology clinics and advertisements. Healthy controls were age- and sex-matched to the PD groups.

All participants provided written informed consent, and the study was approved by the institutional ethics review boards at the University of Iowa and University of South Dakota. This study was conducted in accordance with the Declaration of Helsinki, ensuring confidentiality and voluntary participation. Inclusion criteria for participants who had PD included a clinical diagnosis of idiopathic PD based on the United Kingdom Parkinson’s Disease Society Brain Bank criteria. Exclusion criteria included a diagnosis of dementia, other neurological or psychiatric disorders, or musculoskeletal conditions affecting gait. Healthy controls were required to have no history of neurological or psychiatric illness. Clinical assessments for participants with PD included the motor section of the MDS-Unified Parkinson’s Disease Rating Scale (MDS-UPDRS Part III), to evaluate motor symptom severity, and the FOGQ, to measure freezing severity. Cognitive function was assessed using the Montreal Cognitive Assessment (MOCA). Participant demographics can clinical characteristics are presented in Table 1.

**Table 1.** Demographics and clinical characteristic measurements.

|  | **Control**<br>**(n = 21)** | **PD**<br>**(n = 50)** | **PDFOG–**<br>**(n = 16)** | **PDFOG+(*Normal*)**<br>**(n = 21)** | **PDFOG+ (*Slow*)**<br>**(n = 13)** |
| --- | --- | --- | --- | --- | --- |
| Age (years) | 69.1 ± 1.4 | 68.3 ± 1.0 | 67.3 ± 1.8 | 69.9 ± 1.4 | 67.0 ± 2.4 |
| DD (years) | — | 6.0 ± 0.6 | 5.1 ± 0.7 | 6.0 ± 1.1 | 7.1 ± 1.0 |
| LEDD (mg) | — | 757.1 ± 66.0 | 648.1 ± 100.6 | 750.7 ± 109.7 | 901.5 ± 132.0 |
| MOCA | — | 24.3 ± 0.6 | 26.2 ± 0.6 | 24.0 ± 0.9 | 22.6 ± 1.3 |
| MDS-UPDRS<br>Part III | — | 21.6 ± 2.1 | 12.8 ± 2.1 | 29.3 ± 4.1** | 19.8 ± 1.6 |
| FOGQ | — | 8.9 ± 0.8 | 2.9 ± 0.4 | 10.8 ± 1.1** | 13.2 ± 0.7 |

Values: mean ± SEM
DD, Disease duration; LEDD, Levodopa equivalent daily dose;
MOCA, Montreal Cognitive Assessment; mUPDRS, motor Unified Parkinson's Disease Rating Scale;
FOGQ, Freezing of Gait Questionnaire.
Independent t-test (PDFOG– versus PDFOG+ *normal*): \*p < 0.05; \*\*p < 0.001.
Normal and slow represent the speed of the GI and GO tasks.

### GI and GO tasks

Participants completed two experimental tasks designed to activate gait-related neural activity without requiring physical movement. During the GI task, participants were instructed to vividly imagine themselves walking on a level, unobstructed path, at their natural walking pace. They were asked to visualize the sensation and effort of walking as though they were physically performing the movement. In the GO task, participants watched a video of a computer-generated avatar walking across a flat surface.

Healthy controls and participants in the PDFOG– group completed both tasks at a normal walking speed. Participants in the PDFOG+ group were divided into two subgroups: one performed both tasks at a normal speed, while the other completed both tasks at an intentionally reduced speed. Tasks were presented in randomized order, and all participants received standardized instructions and training to ensure task comprehension and adherence. Participants were instructed to refrain from any overt motor activity during the tasks.

### EEG data acquisition and preprocessing

Both experimental tasks were conducted while participants sat comfortably in a dimly lit, sound-attenuated room. EEG signals were recorded throughout the duration of the tasks. Initially, EEG signals were collected during eyes-closed (2-3 min) and eyes-open (2-3 min) and then GI and GO tasks were performed.

EEG signals were recorded using a 64-channel cap configured according to the international 10–20 system and a Brain Vision amplifier. The sampling rate was set at 500 Hz, and electrode impedances were maintained below 5 kΩ to ensure high signal quality. The reference electrode was placed at Pz, and the ground electrode at Fpz. EEG data preprocessing was performed using MATLAB and the EEGLAB toolbox.^22^ The preprocessing pipeline began with a bandpass filter applied between 0.5 and 50 Hz to remove low-frequency drifts and high-frequency noise. EEG data corresponding to the eyes-closed, eyes-open, normal GI, and normal GO tasks were segmented into 4-second epochs. In contrast, EEG signals for slower GI, slower GO, and their corresponding eyes-closed and eyes-open conditions were segmented into 8-second epochs to match the longer task durations. Artifact removal was conducted in multiple steps. First, bad epochs were identified and removed using the FASTER algorithm,^23^ and the pop_rejchan function in EEGLAB. Independent Component Analysis was then applied to detect and remove artifacts associated with eye blinks and other non-neural sources. Finally, the signals were re-referenced to the average of all channels to minimize reference bias.

### EEG postprocessing

Spectral analyses were performed on the preprocessed and segmented EEG datasets using the pwelch method. A 1-second time window with 50% overlap in window length was applied for spectral estimation. The relative power (frequency-specific power divided by the total spectral power) was computed across five frequency bands: delta (1–4 Hz), theta (4–7 Hz), alpha (7–13 Hz), beta (13–30 Hz), and gamma (30–50 Hz).

We conducted comparisons between GI and corresponding eyes-closed datasets for all channels and frequency bands within the three participant groups (PDFOG+, PDFOG–, and healthy controls). Topographic plots were generated to observe whether the eyes-closed dataset exhibited similar oscillatory patterns to the GI condition. Similarly, comparisons were performed between GO and corresponding eyes-open datasets for all channels and frequency bands within the three groups. Topographic plots were used to assess whether the eyes-open dataset exhibited similar oscillatory patterns to the GO condition.

To further investigate group differences, we generated topographic maps for GI and GO tasks across all frequency bands, comparing the three groups. Additionally, specific comparisons were made between PDFOG+ participants performing normal and slower GI and GO tasks to evaluate the oscillatory activities across all cortical regions. Notably, based on findings from previous work,^21, 24, 25^ we focused on the primary analyses at the midfrontal region, specifically the midfrontal ‘Cz’ electrode. This region of interest was chosen due to its critical role in cognitive control and motor planning, which are believed to be central to gait-related dysfunction in PD, particularly in PDFOG+.^2, 15^

### Statistical analysis

Statistical analyses were performed using MATLAB statistical toolbox and SPSS software. Initially, paired t-tests were conducted to compare resting-state eyes-closed EEG data with GI data for all electrodes and across all five frequency (delta, theta, alpha, beta, and gamma) bands within each group. Multiple comparison corrections were not applied at this stage, and the outcomes were visualized using topographic plots. A similar analysis was performed for the GO task, where resting-state eyes-open data were compared with GO task data. Next, group comparisons were conducted using independent t-tests to assess differences in GI data between the groups (PDFOG+, PDFOG−, and healthy controls) across all electrodes and frequency bands. To evaluate subgroup differences within the PDFOG+ group, independent t-tests were used to compare normal GI with slower GI conditions. The same approach was applied to the GO task, with group comparisons performed between PDFOG+, PDFOG−, healthy controls, and PDFOG+ subgroups. These results were presented using topographic plots to illustrate spatial differences in oscillatory activity.

In the next stage of analysis, we focused on our primary hypothesis regarding midfrontal oscillations, particularly at the ‘Cz’ electrode. Group differences in relative power across each frequency band were assessed using one-way ANOVA, with group (PDFOG+, PDFOG−, and healthy controls) as the between-subject factor. Post-hoc comparisons were conducted using Fisher’s least significant difference procedure to identify significant pairwise differences. Additionally, midfrontal oscillations were compared between the PDFOG+ subgroups (normal versus slower GI/GO) to explore subgroup-specific differences in the frequency bands.

Furthermore, we conducted two-way mixed ANOVAs to examine midfrontal activity across frequency bands, using task type (GI and GO) as the within-subject factor and group as the between-subject factor. Post hoc multiple comparisons were performed using Tukey’s HSD test. Additionally, paired t-tests were used to compare GI-slow and GO-slow conditions within each frequency band, for PDFOG+ subjects.

## Results

### Clinical characteristics measurements

Demographic and clinical comparisons are summarized in Table 1. When contrasting the demographic and clinical characteristics of healthy controls and people with PD, no statistically significant difference in age between the two groups were identified (p >0.05). A comparison between PDFOG+ normal subgroup and PDFOG− group, however, revealed MDS-UPDRS Part III scores which exhibited a difference (p = 0.002), indicating variations in motor symptoms. Additionally, there was a significant difference in performance on the FOGQ between the same groups (p < 0.001). No significant differences were observed between these groups in age (p = 0.26), disease duration (p = 0.51), levodopa equivalent daily dose (LEDD; p = 0.51), or MOCA scores (p = 0.06). However, when comparing the slow- and normal-PDFOG+ subgroups, no statistically significant differences were noted in age (p = 0.27), DD (p = 0.53), LEDD (p = 0.39), or any of the clinical tests, including the MOCA (p = 0.37), mUPDRS (p = 0.88), or the FOGQ (p = 0.11).

### GI versus eyes closed

Prior to the GI task, baseline cortical activity was recorded during an eyes-closed resting state. Oscillatory analysis confirmed expected differences in cortical activity between eyes-closed resting state and the subsequent GI task across multiple frequency bands, channels, and participant groups (Figure 1). These baseline recordings support the idea that these two conditions engage distinct neural processes and provide a reference for interpreting neural activity changes observed during GI.

**Figure. 1.**
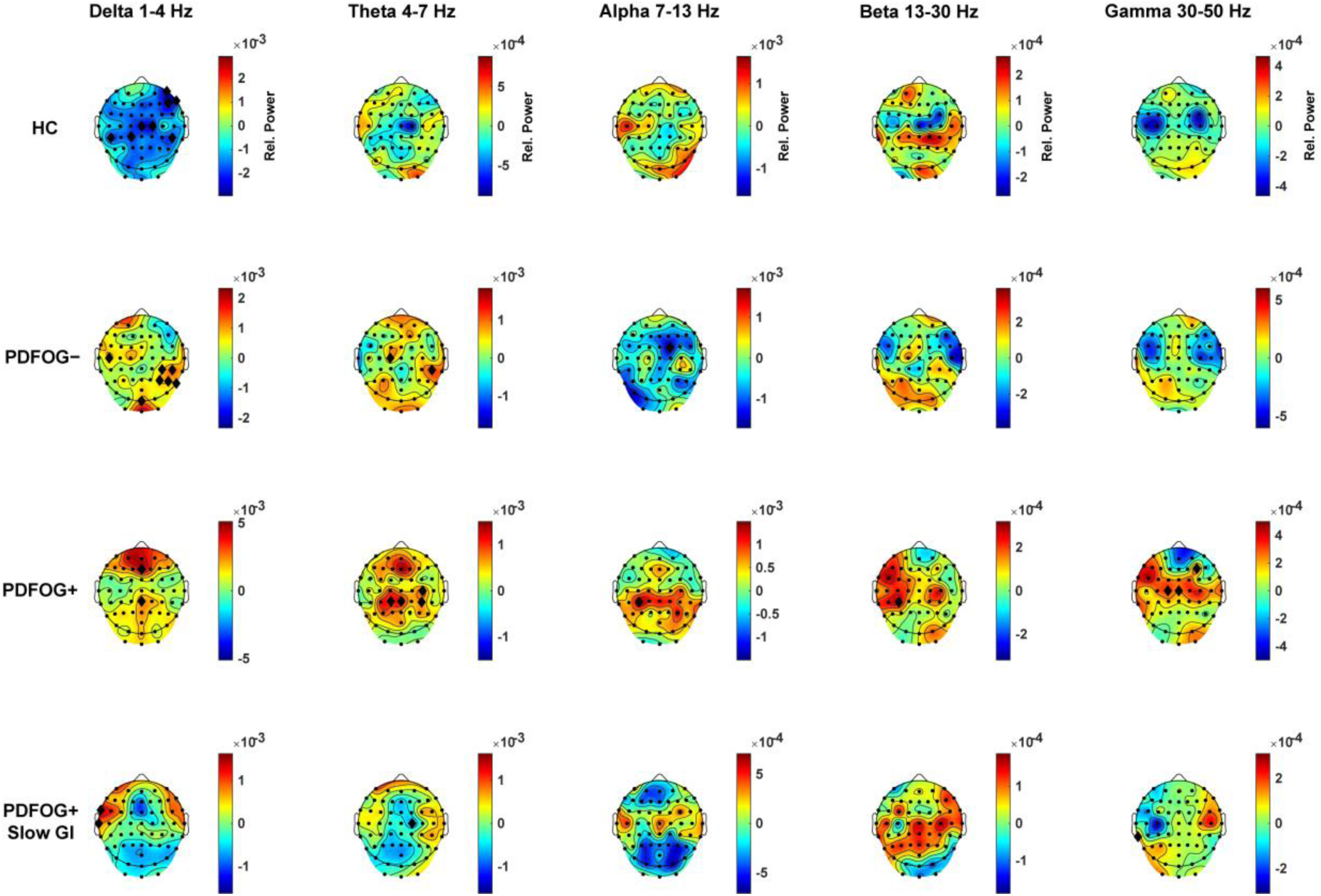
Topographic distribution of oscillatory activity during gait imagination versus eyes-closed resting state. Topographic maps represent the difference in neural oscillatory power between gait imagination and resting with eyes closed, across five frequency bands and all EEG channels. Data are presented for healthy controls (HC), PDFOG–, and PDFOG+. For PDFOG+ subjects, an additional condition with slower imagined gait speed is included. Subtraction of the eyes closed resting state from gait imagination highlights task-related changes in cortical activity. Frequency- and region-specific differences suggest distinct neural dynamics associated with gait imagery across subject groups and conditions. Black diamonds indicate uncorrected statistically significant differences at p < 0.05.

### GI-related midfrontal oscillatory activities in all three groups

During the GI task, significant group differences were observed across frequency bands and channels including the midfrontal cortical region (Figure 2A and Figure S1A). In the delta band, a main effect of group was found (F(2, 55) = 3.85, p = 0.027; Figure 2B). Post-hoc analysis indicated that participants in the PDFOG+ group exhibited significantly higher delta power compared to PDFOG− and healthy control groups. For the theta band, significant differences were found (F(2,55) = 4.15, p = 0.021; Figure 2C), with both PDFOG+ and PDFOG− groups showing increased theta power compared to healthy controls. This suggests that both PD subgroups exhibited enhanced engagement in midfrontal theta oscillations during GI, but there were no significant differences between the two PD groups. The alpha band did not show any significant group differences (F(2,55) = 1.28, p = 0.29; Figure 2D), indicating that this frequency range may not play a critical role in distinguishing neural activity during GI condition. In contrast, the beta band showed significant group effects (F(2,55) = 3.13, p = 0.05; Figure 2E). Post-hoc comparisons revealed a trend in which individuals in the PDFOG− group exhibited significantly higher beta power compared to healthy controls, but interestingly, the PDFOG+ group did not show the same elevation. Instead, individuals in the PDFOG+ group demonstrated decreased beta power compared to the PDFOG− group, which may reflect modulated motor-related cortical engagement in PDFOG+ subjects. For the gamma band, no statistically significant differences were detected among the groups (F(2,55) = 2.42, p = 0.098; Figure 2F).

**Figure. 2.**
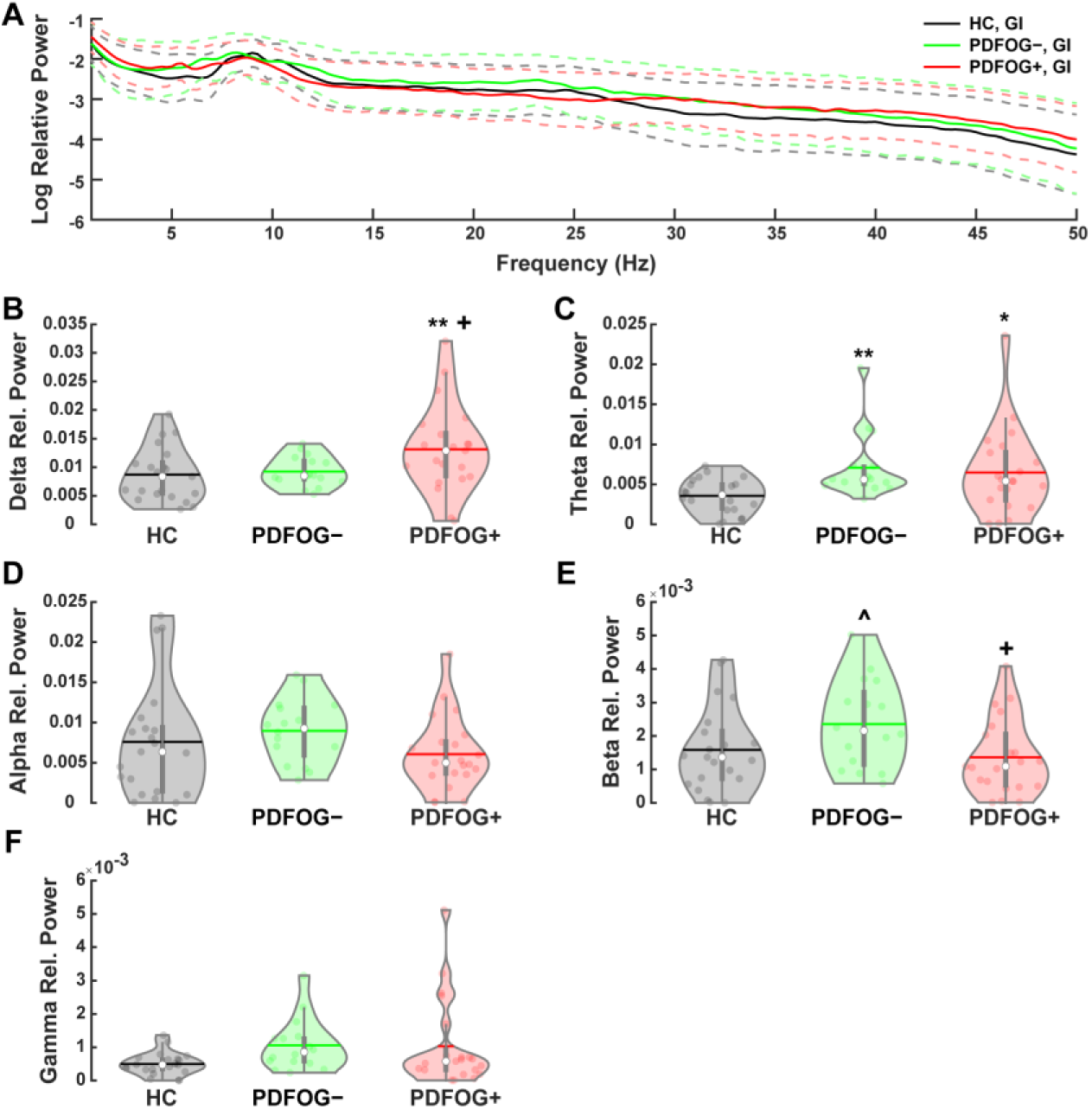
Midfrontal oscillatory activity during gait imagination across groups. (A) Spectral power plots during gait imagination in healthy controls (HC), PDFOG–, and PDFOG+. (B–F) Violin plots showing relative power values in the delta (B), theta (C), alpha (D), beta (E), and gamma (F) frequency bands across the three groups. Dashed lines on the spectral plots indicate the standard error of the mean. Horizontal lines represent mean values; white circles represent medians. p < 0.05 vs. HC (*), p < 0.01 vs. HC (**), p < 0.06 vs. HC (^), p < 0.05 vs. PDFOG– (^+^).

### Effects of GI speed in PDFOG+ subjects

To further investigate the role of GI in motor network activity, PDFOG+ participants were split into two groups and assigned to normal- or slow-GI conditions. The slow-GI subgroup was asked to imagine walking at a reduced pace to reflect their specific impairment. The neural activity of the slow-GI subgroup was compared to the individuals in the normal-GI subgroup. (Figure 3A and Figure S1B). A significant reduction in delta band power was observed in the slow-GI condition compared to normal-paced GI (p < 0.001; Figure 3B). No significant changes in power were noted in the theta, alpha, and beta frequency bands (Figure 3C-E), but we observed a trend with lower power in the gamma band (p = 0.06; Figure 3F) during slow-GI task. Interestingly, decreased power in the delta, theta, beta and gamma bands during slow-GI closely approximated that of the healthy control group, suggesting that imagining gait at a reduced pace may modulate neural activity toward a more normalized pattern.

**Figure. 3.**
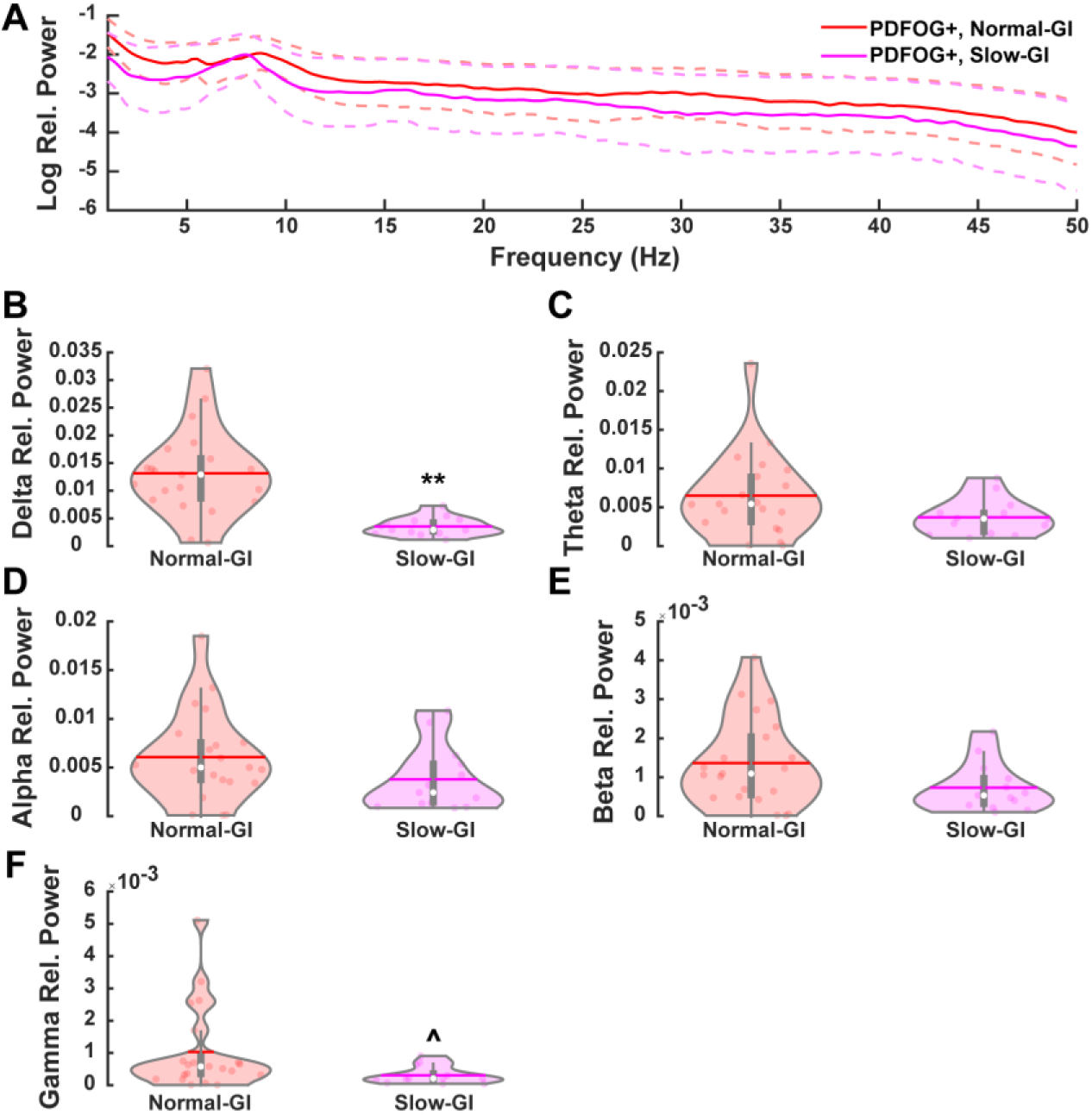
Midfrontal oscillatory activity during normal versus slow gait imagination in PDFOG+ subjects. (A) Spectral power plots comparing normal-speed and slow-speed gait imagination within the PDFOG+ group. (B–F) Violin plots showing relative power values in the delta (B), theta (C), alpha (D), beta (E), and gamma (F) frequency bands under both gait imagination conditions. Dashed lines on the spectral plots indicate the standard error of the mean. Horizontal lines denote mean values; white circles represent medians. p < 0.01 for slow vs. normal gait imagination (**), p < 0.06 (^).

### GO versus eyes open

Prior to the GO task, baseline cortical activity was recorded while participants remained at rest with their eyes open. As presumed, oscillatory activity patterns recorded during eyes open condition differed from those observed during active GO (Figure 4). These baseline measures provided a point of reference for subsequent comparison and, again, support the notion that active observation of gait engages distinct neural processes compared to passive visual perception with eyes open.

**Figure. 4.**
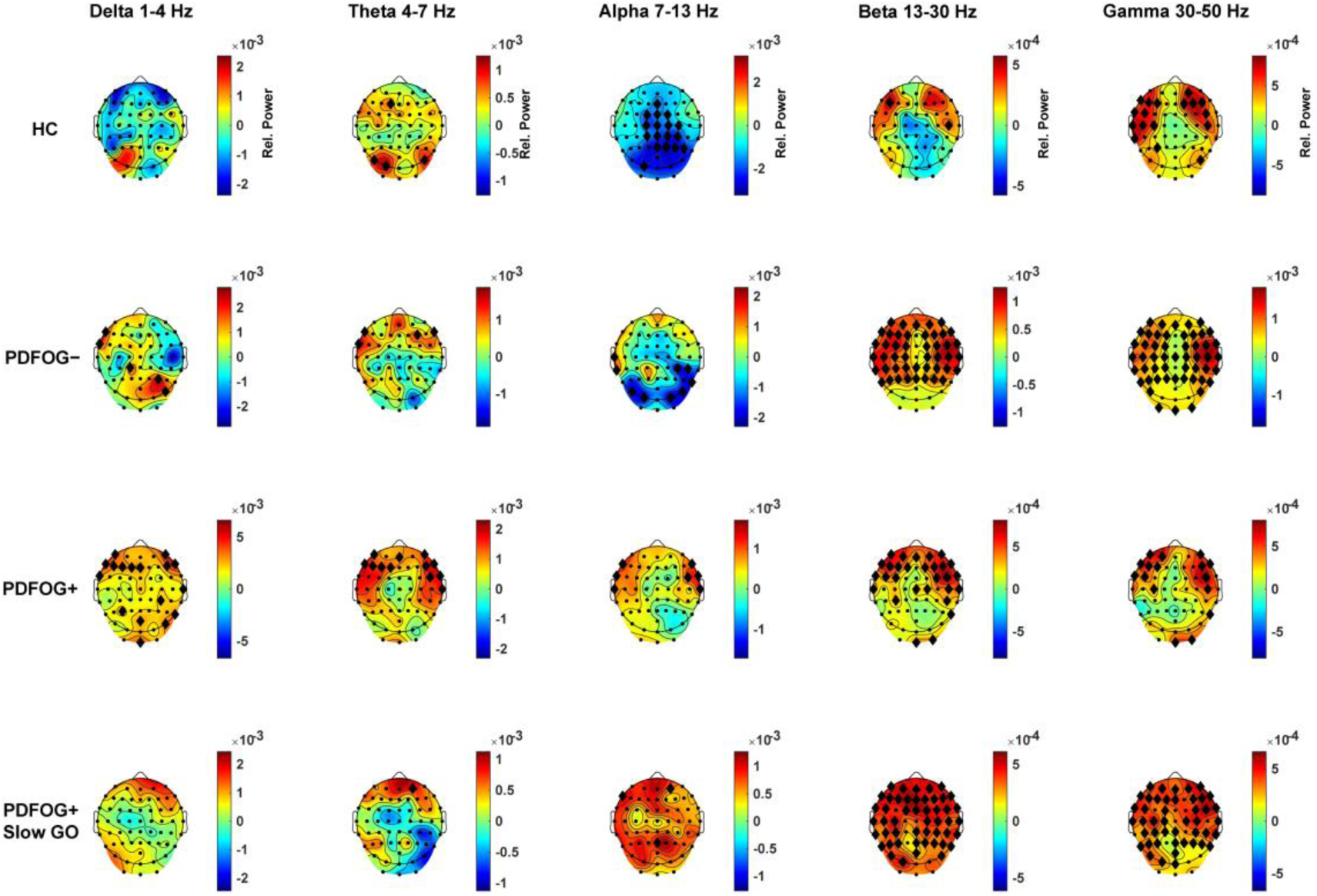
Topographic distribution of oscillatory activity during gait observation versus eyes-open resting state. Topographic maps illustrate the difference in neural oscillatory power between gait observation and eyes-open resting across five frequency bands and all EEG channels. Results are shown for healthy controls (HC), PDFOG–, and PDFOG+. For PDFOG+ subjects, an additional condition involving observation of slower gait is included. Neural activity during gait observation was isolated by subtracting eyes-open resting-state data, revealing task-related modulations. Observed frequency- and region-specific changes suggest altered cortical dynamics during gait observation across different groups and conditions. Black diamonds indicate uncorrected statistically significant differences at p < 0.05.

### GO-related midfrontal oscillatory activities in all three groups

During the GO task, significant group differences were observed across frequency bands and channels including the midfrontal cortical region (Figure 5A and Figure S2A). In the delta band, no significant differences were detected among the three groups (F(2,55) = 2.36, p = 0.104; Figure 5B). In the theta band, a significant difference was observed (F(2,55) = 3.47, p = 0.038; Figure 5C). Post-hoc analysis indicated that the PDFOG− group exhibited significantly greater theta power compared to healthy controls, whereas no significant differences were detected between the PDFOG+ group and healthy controls or between the two PD subgroups. Interestingly, the theta oscillatory activity observed during GO closely resembled that found in GI, suggesting common neural mechanisms underlying these two tasks. Significant differences were also observed in the alpha band (F(2,55) = 4.97, p = 0.01; Figure 5D). Specifically, the PDFOG− group demonstrated higher alpha power compared to both healthy controls and the PDFOG+ group, while no significant differences were found between the PDFOG+ group and healthy controls. In the beta band, a significant group effect was also identified (F(2, 55) = 3.52, p = 0.037; Figure 5E). The PDFOG− group exhibited greater beta power compared to both healthy controls and the PDFOG+ group, whereas the PDFOG+ group showed significantly lower beta power relative to those in the PDFOG− group. Notably, no significant differences were found between the PDFOG+ group and healthy controls, mirroring the pattern observed in GI. A trend toward significance was found in the gamma band (F(2,55 = 2.94, p = 0.06; Figure 5F). Both the PDFOG+ and PDFOG− groups demonstrated increased gamma power relative to healthy controls, but no significant differences were found between the PD subgroups.

**Figure. 5.**
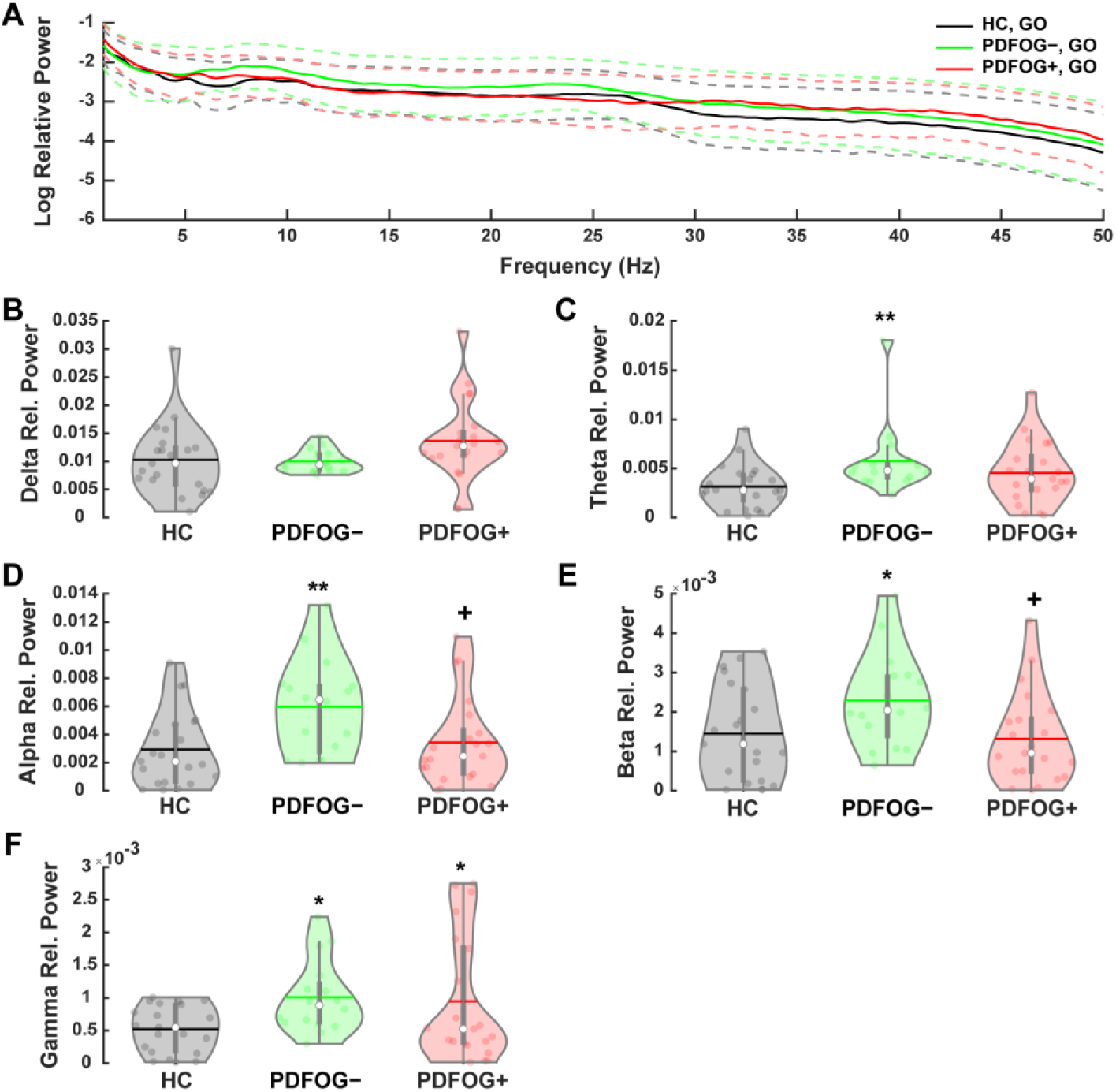
Midfrontal oscillatory activity during gait observation across groups. (A) Spectral power plots during gait observation in healthy controls (HC), PDFOG–, and PDFOG+. (B–F) Violin plots showing relative power values in the delta (B), theta (C), alpha (D), beta (E), and gamma (F) frequency bands across the three groups. Dashed lines on the spectral plots indicate the standard error of the mean. Horizontal lines denote mean values; white circles represent medians. p < 0.05 vs. HC (*), p < 0.01 vs. HC (**), p < 0.05 vs. PDFOG– (^+^).

### Effects of GO speed in PDFOG+ subjects

To investigate the impact of observed gait speed on midfrontal oscillatory activity, the PDFOG+ participants were divided and assigned to a slow- or normal-speed GO task. Those who were assigned to the slow-GO task were asked to watch an avatar walking at a reduced pace, to mimic their own impairments. Those in the normal-GO task subgroup watched an avatar walking at a normal speed. Cortical activity was monitored during this task in both groups and revealed some significant differences (Figure 6A and Figure S2B). Compared to those in the normal-paced GO condition, the PDFOG+ participants in the slow-GO condition demonstrated significant reductions in delta band (p = <0.001; Figure 6B) and theta band (p = 0.01; Figure 5C) power. No significant changes in power were noted in the alpha, beta, and gamma frequency bands (Figure 6D-F). Interestingly, similar to slow-GI task, the oscillatory activity recorded during the slow-GO condition closely resembled that of healthy controls, suggesting that observing gait at a slower pace may normalize cortical activity in individuals in this PDFOG+ subgroup.

**Figure. 6.**
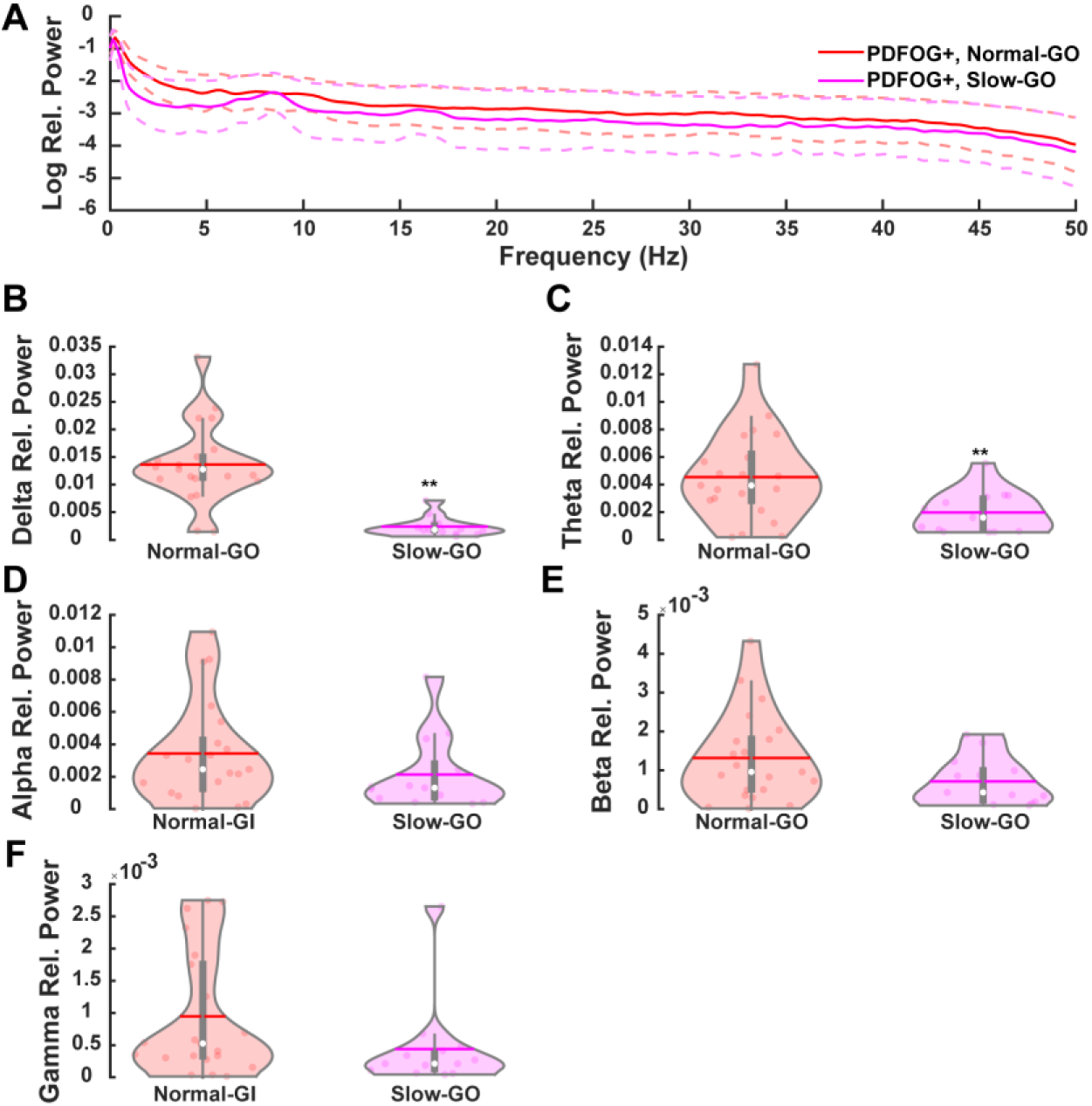
Midfrontal oscillatory activity during normal versus slow gait observation in PDFOG+ subjects. (A) Spectral power plots comparing observation of normal-speed versus slow-speed gait within the PDFOG+ group. (B–F) Violin plots showing relative power values in the delta (B), theta (C), alpha (D), beta (E), and gamma (F) frequency bands across both observation conditions. Dashed lines on the spectral plots indicate the standard error of the mean. Horizontal lines denote mean values; white circles represent medians. p < 0.01 for slow vs. normal gait observation (**).

### Comparison of GI and GO tasks

Table 2 shows the direct comparison of the oscillatory patterns associated with GI and GO tasks. Significant within-subject effects were observed in the theta (F(1, 55) = 12.98, p <0.001) and alpha (F(1,55) = 40.51, p < 0.001) bands. No significant within-subject differences were detected in the delta, beta or gamma bands. Between-subject effects revealed group differences in delta (F(2,55) = 3.51, p = 0.037), theta (F(2, 55) = 4.22, p = 0.02), and beta (F(2,55) = = 3.57, p = 0.03) frequencies. Post hoc comparison showed that theta activity significantly differed between healthy control and PDFOG– groups (p = 0.021), and beta activity differed between PDFOG– and PDFOG+ groups (p = 0.035). No significant interaction effects were observed in any other frequency band. Furthermore, paired t-tests supported the within-subject ANOVA findings, with significant GO-slow versus GI-slow differences for theta (t(12) = 3.87, p = 0.002) and alpha (t(12) = 2.40, p = 0.034) frequencies.

**Table 2.** Summary of two-way mixed ANOVAs and paired t-tests.

|  | **Delta band** | **Theta band** | **Alpha band** | **Beta band** | **Gamma band** |
| --- | --- | --- | --- | --- | --- |
| **Within-subject effect**<br>(GO vs. GI) | F(1, 55) = 2.69,<br>p = .11 | F(1, 55) = 12.98,<br>p < .001* | F(1, 55) = 40.51,<br>p < .001* | F(1, 55) = 0.89,<br>p = .350 | F(1, 55) = 0.24,<br>p = .62 |
| **Between-subject effect**<br>(HC, PDFOG–, PDFOG+) | F(2, 55) = 3.51,<br>p = .037* | F(2, 55) = 4.22,<br>p = .020* | F(2, 55) = 2.32,<br>p = .11 | F(2, 55) = 3.57,<br>p = .035* | F(2, 55) = 2.94,<br>p = .06 |
| **Interaction effect**<br>(GO vs. GI x Group) | F(2, 55) = 0.34,<br>p = .71 | F(2, 55) = 1.88,<br>p = .16 | F(2, 55) = 1.45,<br>p = .24 | F(2, 55) = 0.11,<br>p = .90 | F(2, 55) = 0.20,<br>p = .820 |
| **Post hoc comparisons**<br>(p-values) | p<sup>12</sup> = .99<br>p<sup>13</sup> = .053<br>p<sup>23</sup> = .09 | p<sup>12</sup> = .02*<br>p<sup>13</sup> = .10<br>p<sup>23</sup> = .69 | p<sup>12</sup> = .23<br>p<sup>13</sup> = .91<br>p<sup>23</sup> = .11 | p<sup>12</sup> = .10<br>p<sup>13</sup> = .87<br>p<sup>23</sup> = .035* | p<sup>12</sup> = .10<br>p<sup>13</sup> = .11<br>p<sup>23</sup> = .98 |
| **Paired t-test**<br>(GO-slow vs. GI-slow) | t(12) = 1.79,<br>p = .098 | t(12) = 3.87,<br>p = .002* | t(12) = 2.40,<br>p = .034* | t(12) = 0.12,<br>p = .90 | t(12) = –0.86,<br>p = .40 |

p<sup>12</sup> = Healthy controls versus PDFOG–; p<sup>13</sup> = Healthy controls versus PDFOG+; p<sup>23</sup> = PDFOG– versus PDFOG+.
Statistically significant at \*p < 0.05.

## Discussion

Our study demonstrates that slowing the pace of GI and GO in people with PDFOG+ can partially restore midfrontal oscillatory activity toward patterns seen in healthy controls. This normalization likely arises from reduced cognitive-motor demands, enabling executive control networks to function more efficiently and helping to prevent the timing and planning disruptions that lead to freezing episodes. By easing temporal pressure, slow-paced tasks likely facilitate compensatory recruitment of frontal and parietal regions, improving cognitive-motor integration and rebalancing cortical and brainstem contributions to gait control. The distinct neural signatures observed for GI and GO highlight that each engages partially separate circuits. GI appears to emphasize internally driven motor planning and GO relies on externally triggered perception-action coupling. Both of these circuits are likely impaired in PDFOG+. These finding support the view that FOG reflects a network-level failure in both automatic and cognitively mediated gait control, and that modifying task parameters can leverage residual neuroplasticity to recalibrate dysfunctional circuits.

### Mechanisms of oscillatory normalization in slow-paced tasks

Our findings suggest that when people with PD and FOG perform GI or GO at a slowed pace, midfrontal oscillatory activity shifts toward patterns seen in healthy controls. Slower pacing reduces the timing pressure and cognitive load required for gait simulation, which may prevent the overtaxing of executive resources that contribute to FOG episodes.^12, 26^ Behavioral studies have shown that applying attentional strategies or external pacing cues can alleviate FOG by bypassing impaired internal timing circuits.^27, 28^ For example, consciously modulating gait or using rhythmic cues engages alternative pathways and often reduces freezing frequency in people with PD.^12, 27, 28^ Similarly, slowing the imagined or observed gait provides a form of “external” temporal structure or additional processing time that helps people with PD recruit compensatory networks more effectively, thereby partially restoring normal oscillatory patterns.

Physiologically, our study demonstrated that the midfrontal cortex appears to retain plasticity in PD, such that adjusting task parameters can modulate the oscillatory activity in these regions. During slow-GI, individuals in the PDFOG+ group demonstrated a significant reduction in midfrontal delta power relative to normal-paced conditions, bringing delta activity closer to healthy control levels, and suggesting a potential normalization of cortical timing mechanisms under reduced task demands. Although no statistically significant differences were observed in theta, alpha, or beta bands during slow-GI, the significant change in delta pattern and a trend toward reduced gamma activity are consistent with evidence that PDFOG+ exhibit impaired midfrontal delta oscillations and interval timing.^29, 30^

### Cognitive-motor integration and frontal compensatory networks

The normalization of EEG rhythms in slow GI and GO likely reflects more effective cognitive-motor integration, when task difficulty is minimized. FOG is increasingly viewed as a network-level failure wherein both motor automaticity and cognitive control are compromised.^31, 32^ In healthy gait, higher cortical centers provide seamless top-down regulation of brainstem and spinal pattern generators.^31, 33^ In PD, dopamine loss and basal ganglia dysfunction weaken these automatic pathways, forcing greater reliance on conscious, frontal lobe strategies.^31^ This compensation is limited in PDFOG+, who often experience signs of executive dysfunction and diminished conflict-resolution capacity, especially under dual-task or high-demand conditions, manifesting in aberrant midfrontal oscillations.^32^

Slower-paced GI and GO mitigate these issues by lowering cognitive load, allowing frontal executive networks to engage more efficiently without being overwhelmed.^29^ Our result that midfrontal theta activity increased toward normal in the PDFOG+ group under the slow-GO condition supports this interpretation, and is consistent with the role of theta oscillations in cognitive control and conflict monitoring.^16, 31^ By facilitating an increase in theta, slower GI/GO may enable better conflict resolution and plan-updating, shifting gait control from a failure-prone automatic mechanism (dominated by basal ganglia loops) to a compensatory mode relying on frontal networks. Imaging studies similarly show that people with PD, especially those with FOG, recruit extensive frontal and parietal regions when imagining or planning complex gait actions, far more than controls.^14, 34, 35^ This broader cortical recruitment represents a compensatory mechanism to overcome deficient automatic gait generation and underscores that the integration of cognitive and motor processes is critical to maintaining locomotion.

In this study, when we slowed down the imagined or observed gait for the PDFOG+ group, they may have effectively tapped into these compensatory cortical resources. Slowing the task likely synchronized the timing between cognition and motor imagery, giving frontal executive areas sufficient time to formulate a “motor plan” and relay it to motor circuits without the usual processing speed demands that could induce errors or freezing. By easing task demands, slow GI or GO might allow frontal cortical input to be more effective and reduce the need for accelerated basal ganglia and brainstem output, thereby preventing the network breakdown that triggers freezing.

### GO versus GI: distinct neural circuits

Although both GI and GO engage overlapping motor networks, they recruit partially distinct neural circuits. GI is an internally driven process that strongly involves motor planning regions such as the supplementary motor area (SMA), pre-SMA, dorsolateral prefrontal cortex (DLPFC), and motor cortices.^9^ During motor imagery of gait, healthy individuals activate much of the same cortical-subcortical networks used for actual movement execution.^9^ Alternatively, GO relies on externally triggered perceptual-motor coupling via the mirror neuron system.^12, 13^ Activation of this system involves visual motion-processing areas and the parieto-frontal mirror network, including the premotor cortex and inferior parietal lobule, and can subliminally recruit the motor circuits of observer corresponding to the observed actions.^9, 12^ These differences carry implications for people with PD who experience FOG. GO tasks might be cognitively less demanding than GI because an external visual stimulus guides the simulation and its efficacy may depend on mechanisms that link perception to action. Consistent with this distinction, our midfrontal EEG data identified GO and GI differences in the theta and alpha bands, suggesting that the two tasks recruit partially distinct control strategies. Prior fMRI studies also report reduced activation during action observation of normal walking in frontal, cingulate, and parieto-occipital regions in people with FOG, with larger reductions in PDFOG+ than in PDFOG– or healthy controls.^13^ In our study, beta power differed between PDFOG+ and PDFOG– across tasks. Given the absence of a task-by-group interaction, we do not ascribe this effect specifically to GO. We interpret it as a task-general alteration in midfrontal control that is compatible with action-observation hypoactivation reported in the literature, while noting that our measures are restricted to midfrontal oscillations and do not index posterior mirror-network contributions. This pattern could reflect reduced sensorimotor resonance, but we cannot confirm the underlying sources with our data. Conversely, GI forces engagement of motor schemas and internal cueing. Although this can be challenging in PD, it may directly train the weakened internal route. Clinically, individuals with FOG often rely on external cues, so GO may provide a workable entry point, with GI introduced to strengthen internal generation over time.^13, 26^

### Clinical implications and future directions

Our findings carry several implications for developing interventions to manage FOG and more broadly enhance gait in people with PD. First, they provide neurophysiological support for motor imagery training as a therapeutic tool. Mental rehearsal of walking has been shown to activate similar neural circuits as actual gait and can strengthen these pathways to enhance gait outcomes.^30^ Our results suggest that such imagery should perhaps be done at a comfortable or consciously slower pace for PDFOG+, at least initially. By tailoring imagery speed to each participant’s capacity, we might maximize midfrontal engagement without provoking frustration or neural “overload.” This could gradually entrain the frontal executive-motor pathways to become more efficient.

Similarly, action observation treatment may hold promise as an adjunctive intervention to GI. A previous study reported that a short course of action observation training significantly reduced FOG episodes in PDFOG+.^12^ There is also evidence that combining action observation with simultaneous imagery yields greater cortical activation than either alone, which might amplify the rehabilitative effect.^35, 36^ Our results showing distinct but complementary benefits of GO and GI lend support to this approach.

Another implication is the utility of external cuing strategies for normalizing oscillations and improving gait. Rhythmic auditory stimulation or visual cues are well-established aids that help bypass impaired internal timing in PD.^28^ From an oscillatory perspective, external cues may work by entraining cortical rhythms to the cue frequency, effectively replacing the missing internal rhythm. For instance, auditory cuing tends to reduce excessive beta synchronization and enhance connectivity between auditory, visual, and motor areas.^35^ Our findings with slow paced GI can be seen as an internalized form of cueing. Under this condition the participant’s intentional slow tempo acts as a “cue” that their cortex can more easily follow. Clinicians might leverage this by training patients to adopt a slower, rhythmic counting or humming while initiating gait or during turns, essentially self-cuing to engage frontal control. New technologies like wearable vibrotactile metronomes have also been explored and could be combined with imagery training to reinforce these oscillatory patterns in everyday walking.

Lastly, our study points toward potential roles for non-invasive brain stimulation in treating individuals with FOG. Given the centrality of midfrontal regions in related oscillatory abnormalities, targeting these areas with transcranial stimulation could modulate dysfunctional rhythms. For example, transcranial alternating current stimulation at theta frequency over frontal midline might enhance endogenous theta oscillations and improve cognitive control during gait. Similarly transcranial direct current stimulation over the supplementary motor area or dorsolateral prefrontal cortex has shown preliminary success in improving gait and executive function, in people with PD, by presumably boosting the underlying network excitability.^37, 38^ Deep brain stimulation (DBS) of the subthalamic nucleus, while primarily addressing motor symptoms, has also been observed to shorten beta bursts and reduce freezing severity in some people with PD, especially when using lower-frequency stimulation paradigms.^20^ Future interventions could explore adaptive DBS that responds to beta or theta activity in real-time, using changes in oscillations as a stimulus for adjusting brain stimulation parameters. While such approaches are promising, our results underscore the value of these oscillatory markers, which could serve as feedback signals for closed-loop therapies or as outcome measures to test the efficacy of interventions like cognitive training or cuing.

## Data Availability

All data produced in the present study are available upon reasonable request to the corresponding author.

## Acknowledgements

The authors would like to sincerely thank all research assistants who contributed to data collection for this study.

## Author Contributions

Matthew Leedom: Conceptualization; Formal analysis; Methodology; Supervision; Writing— original draft; and Writing—review & editing. Arturo I. Espinoza: Conceptualization; Data curation; Formal analysis; Investigation; Methodology; and Writing—review & editing. Martina Mancini: Methodology; and Writing—review & editing. Daniel H. Lench: Methodology; and Writing—review & editing. Arun Singh: Conceptualization; Data curation; Formal analysis; Investigation; Methodology; Supervision; Writing—original draft; and Writing—review & editing.

