## Supplementary Figures 1 and 2 for "Midfrontal Oscillatory Alterations During Gait Imagination and Observation in Parkinson’s Disease with Freezing of Gait"

### Supplementary Materials

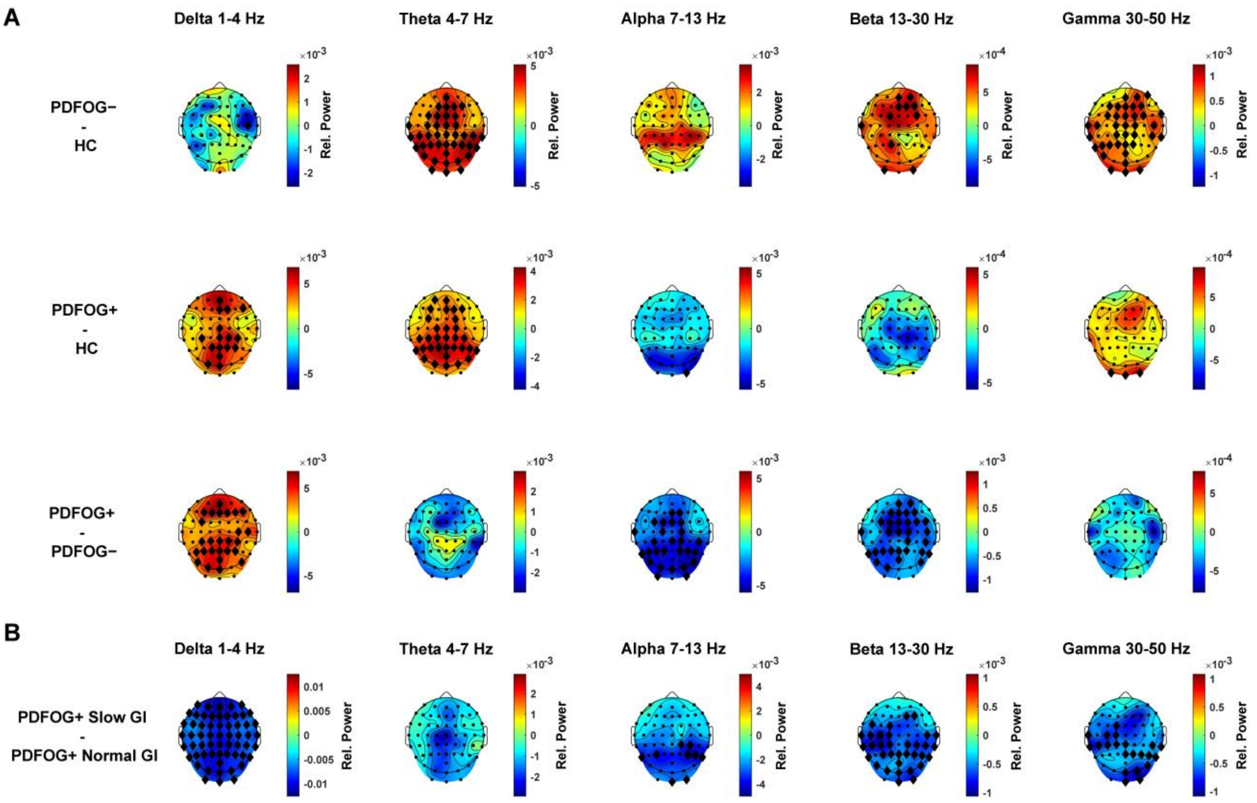


**Figure S1. Topographic distribution of oscillatory activity during gait imagination: between-group comparisons.** Topographic maps illustrate differences in neural oscillatory power during the gait imagination task across five frequency bands and all EEG channels. (A) Comparisons are shown between PDFOG– and healthy controls (HC), PDFOG+ and HC, and PDFOG+ and PDFOG–. (B) Comparisons between PDFOG+ subgroups are presented for gait imagination at slower versus normal imagined gait speeds. Black diamonds indicate uncorrected statistically significant differences at $p < 0.05$.

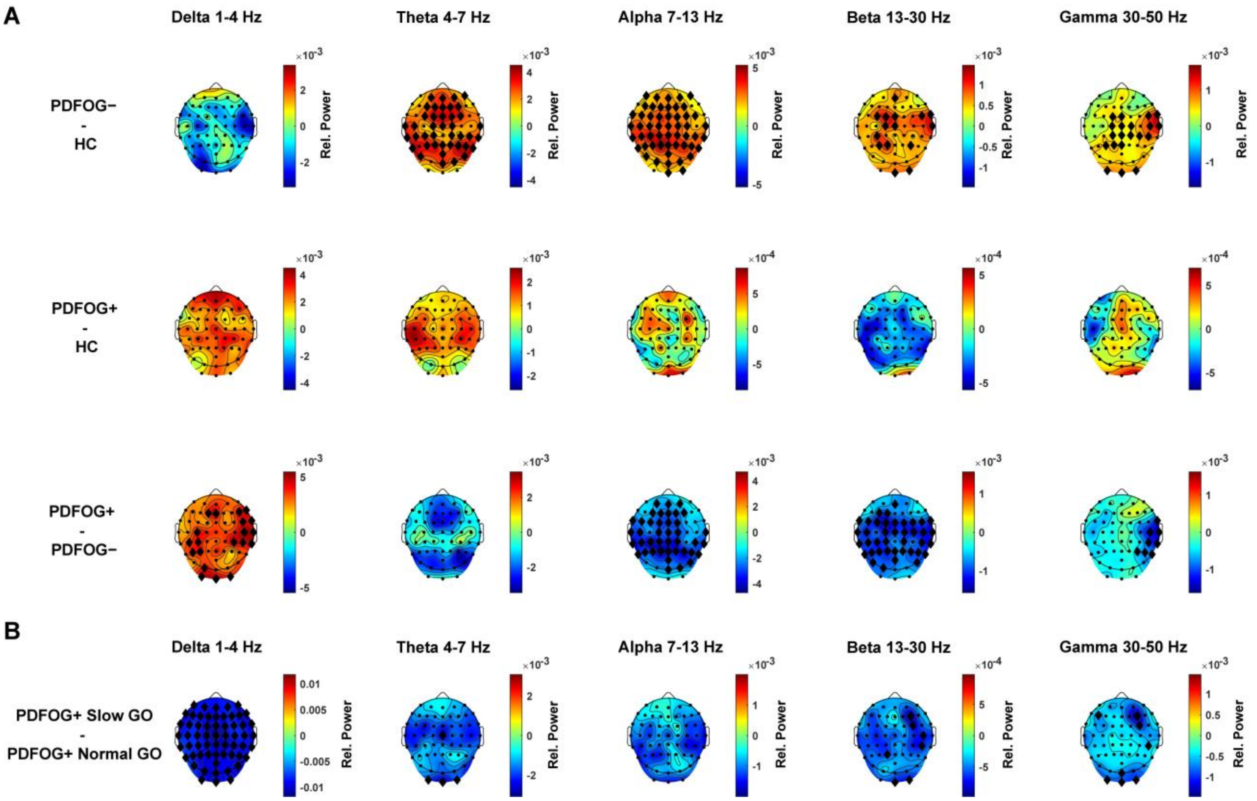


**Figure S2. Topographic distribution of oscillatory activity during gait observation: between-group comparisons.** Topographic maps illustrate differences in neural oscillatory power during the gait observation task across five frequency bands and all EEG channels. (A) Comparisons are shown between PDFOG– and healthy controls (HC), PDFOG+ and HC, and PDFOG+ and PDFOG–. (B) Comparisons between PDFOG+ subgroups are presented for observation of slower versus normal gait speeds. Black diamonds indicate uncorrected statistically significant differences at $p < 0.05$.
